# Application of qualifying variants for genomic analysis

**DOI:** 10.1101/2025.05.09.25324975

**Authors:** Dylan Lawless, Ali Saadat, Mariam Ait Oumelloul, Simon Boutry, Veronika Stadler, Sabine Österle, Jan Armida, David Haerry, D. Sean Froese, Luregn J. Schlapbach, Jacques Fellay

## Abstract

**Motivation:** Qualifying variants (QVs) are genomic alterations selected by defined criteria within analysis pipelines. Although crucial for both research and clinical diagnostics, QVs are often seen as simple filters rather than dynamic elements that influence the entire workflow. In practice these rules are embedded within pipelines, which hinders transparency, audit, and reuse across tools. A unified, portable specification for QV criteria is needed.

**Results:** Our aim is to embed the concept of a “QV” into the genomic analysis vernacular, moving beyond its treatment as a single filtering step. By decoupling QV criteria from pipeline variables and code, the framework enables clearer discussion, application, and reuse. It provides a flexible reference model for integrating QVs into analysis pipelines, improving reproducibility, interpretability, and interdisciplinary communication. Validation across diverse applications confirmed that QV based workflows match conventional methods while offering greater clarity and scalability.

**Availability:** The source code and data are accessible at the Zenodo repository https://doi.org/10.5281/zenodo.17414191. Manuscript files are available at https://github.com/DylanLawless/qvApp2025lawless. The QV framework is available under the MIT licence, and the dataset will be maintained for at least two years following publication.

## 1 Introduction

Qualifying Variant (QV)s are genomic alterations selected by specific criteria within genome processing pipelines, serving as dynamic elements essential for both research and clinical diagnostics. QVs are not merely static filters applied at a single step in an analysis pipeline; rather, they are dynamic, multifaceted elements that permeate the entire workflow, from initial data quality control to final result interpretation. This nuanced perspective underscores that QVs play an integral role in shaping the fidelity and reproducibility of genomic analyses, enabling the iterative refinement of data and facilitating the integration of diverse analytical strategies throughout the pipeline.

Often, QV selection adheres to established variant classification and reporting standards (1–5) and standardised workflows (6–8). However, a unified framework for QVs is lacking, despite the recognised benefits of similar initiatives, such as Polygenic Risk Score (PRS) reporting standards (9; 10). Tools such as vcfexpress (11) enable flexible filtering and formatting of Variant Call Format (VCF) files using user-defined expressions. Treating QV criteria as an external parameter layer complements these tools by externalising their thresholds and logic. This approach improves reproducibility across distributed computing environments (12) and integrates seamlessly with workflow managers like Snakemake (13) or Nextflow (14).

QV selection criteria vary by application. In Genome-Wide Association Study (GWAS), thresholds favour common variants, yielding datasets with over 500,000 variants per subject, whereas rare disease analyses use stringent filters producing fewer than 1,000 variants, often limited to known genes or pathogenic loci. Although targeted filtering is valuable (15; 16), no unified approach exists. In practice, QV sets range from broad quality control filters to specific disease panels, and their definition is critical for reproducibility and accurate reporting, influencing results as much as the pipeline itself (17).

As Whole Genome Sequencing (WGS) becomes standard for large cohorts (18; 19), the integration of diverse QV protocols is critical for data cleaning and analysis. During sequencing analysis several layers can be responsible for triggering QV protocols, including pre-existing metadata, technical Quality Control (QC) results, and post-calling annotations, highlighting the need for a clear, unified approach.

We introduce the QV as a standalone entity, independent from other pipeline variables. Structured human- and machine-readable criteria, aligned with FAIR principles (20), facilitate integration across databases (21; 22). We advocate for the use of standard vocabularies, unique identifiers, and flexible file formats to support this integration.

Building on this framework, we propose an openly documented registry model for QV files that assigns a unique `qv_set_id` and records a SHA-256 checksum for each release, enabling direct retrieval and verification for audit and re-analysis. Our accompanying HTML-based QV builder converts simple `key=value` statements into structured YAML and can be embedded in public, private, or commercial websites to simplify the authoring of consistent criteria (Zenodo repository). The framework is designed to support the emergence of a shared, widely adopted registry over time.

## 2 Methods

### 2.1 Implementation

The QV file provides a structured, human- and machine-readable definition of variant qualifying criteria. It is composed of five logical components that define its structure and metadata. It is portable across tools, transparent in content, and verifiable through unique identifiers and checksums. Each file is a lightweight YAML or JSON document specifying the variables and thresholds used in analysis. It can be read programmatically at runtime, for example using yq in shell-based workflows or `yaml::read_yaml()` in R, providing the same parameters that would otherwise be embedded within pipeline configurations, as illustrated in **Figure 1**. The output is identical to that of the native workflow, with the added benefit of an explicit, versioned, and shareable configuration file.

- **1. Meta**: Descriptive metadata including `qv_set_id`, title, version, author list, creation date, and tags. These fields ensure traceability and version control across analyses.
- **2. Filters**: Simple rule-based statements that apply inclusion or exclusion logic based on variable thresholds (for example, minimum allele frequency or coverage depth). Filters can also restrict the analysis to defined genomic regions, such as a target gene panel or BED file.
- **3.Criteria**: Compound logic blocks that combine one or more conditions into interpretable rules, corresponding to concepts such as ACMG criteria or study-specific thresholds.
- **4. Notes**: Optional free-text annotations providing context, assumptions, or technical caveats.
- **5. Descriptions (optional)**: Plain-language fields, such as `description_patient` and `description_ppie`, that can record patient preferences or public involvement input. These complement the technical definitions without affecting computational logic.

**Figure 1:**
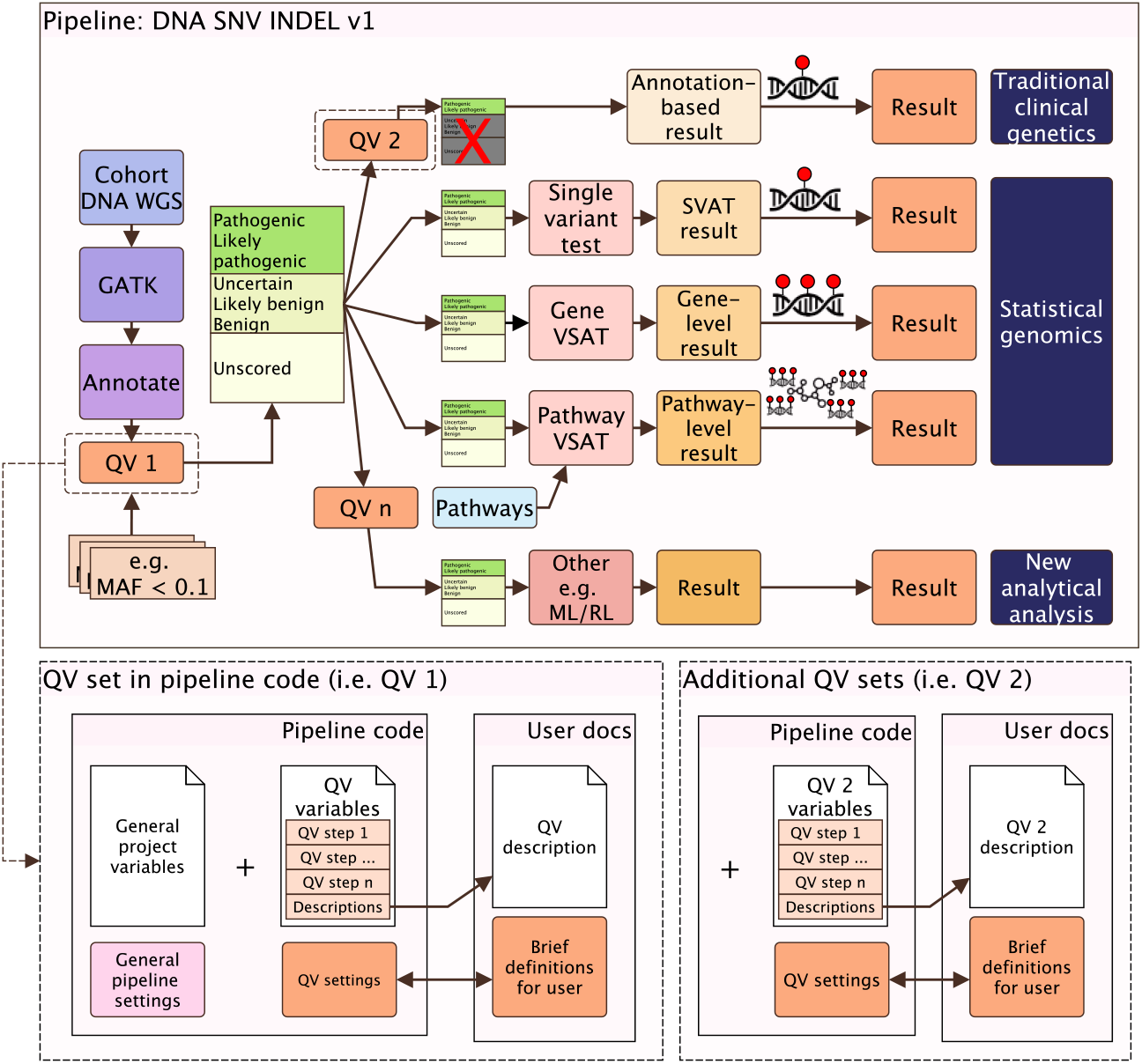
Summary of the QV application for a WGS pipeline. QV1 and QV2 are applied as sequential protocol steps. In this example, QV2 differs from QV1 by retaining only likely/pathogenic variants (indicated by a red X). The QV file loaded by the analysis pipeline comprises a description field (optional) and a variables field (mandatory). The QV criteria may be distributed across multiple pipeline steps.

#### Example QV structure

We include an HTML-based QV builder that can be embedded in research or commercial platforms to simplify the creation of consistent, versioned criteria files (available via Zenodo repository). A minimal QV YAML file is shown in **Box 1** , equivalent to the configuration generated by this builder. QV files are composed of `key=value` statements, ensuring that all filtering and interpretation rules are explicit, versioned, and reproducible. In simple terms, **Box 1** specifies that only variants overlapping a curated disease gene panel are retained and that variants classified as pathogenic or likely pathogenic are prioritised. It also records patient context and patient-public involvement notes, thereby linking the technical filtering logic with its clinical and ethical rationale.

##### Box 1: qv_disease_panel_example.yaml

~~~
meta:
  qv_set_id: qv_disease_panel_v1_20250828
  version: 1.0.0
  title: Disease panel filter
filters:
  region_include:
    description: >
      Restrict to curated disease gene panel
    logic: keep_if
    field: OVERLAP(targets.disease_panel.bed)
    operator: ‘>=‘
    value: 1
criteria:
  pathogenic:
    description: >
      Variant classified as pathogenic or likely pathogenic
    logic: and
    conditions:
      -group: any_of:start
      -{ field: CLASS, operator: ‘==‘, value: P }
      -{ field: CLASS, operator: ‘==‘, value: LP }
      -group: any_of:end
meta:
  description_patient: >
    We have a strong family history of early heart attacks.
  description_ppie: >
    The PPIE group reviewed the criteria and approved them on 2025-08-15.
notes:
  -Gene panel file defines the target regions.
  -Additional quality filters may be added as needed.
~~~

#### FAIR mapping and patient involvement

Each QV file includes a persistent identifier (`qv_set_id`) that links criteria across analyses and databases. The framework aligns with the Findable, Accessible, Interoperable, and Reusable (FAIR) principles of findability, accessibility, interoperability, and reusability (20). Findability is achieved through unique identifiers; accessibility through open, human- and machine-readable YAML or JSON files; interoperability through standardised syntax (i.e. `key=value`) and semantic mappings such as Resource Description Framework (RDF) or Systematized Nomenclature of Medicine-Clinical Terms (SNOMED CT) (21; 22); and reusability through embedded metadata, checksum verification, and versioned registry records.

Optional metadata fields such as `description_patient` and `description_ppie` allow patient input and Public and Patient Involvement and Engagement (PPIE) feedback to be recorded in a manner appropriate to the study or application, with patient notes provided through consent-linked forms and PPIE groups offering structured review or approval of criteria within the same FAIR-compliant file.

#### Example QVs in WGS analysis

A typical WGS pipeline applies several QV sets sequentially, as the genetic cause of disease may stem from different variant types such as SNVs, CNVs, or structural variants. Each pass filters data for its purpose, producing both cohort-level and single-patient results within one reproducible framework (23; 24). As illustrated in **Figure 1**, the description can be written as:

“A cohort of patient WGS data was analysed to identify genetic determinants for phenotype X. A flexible QV set was applied using the `pipeline v1` , which implements the `QV_SNV_INDEL_1` criteria to produce the prepared dataset (`dataset v1`). This dataset was analysed alongside other modules (e.g. `PCA_SNV_INDEL_v1` and `statistical_genomics_v1`) to derive a cohort-level association signal (Result 1). It was then re-filtered with stricter `QV_SNV_INDEL_2` criteria to identify known causal variants, yielding (`dataset v2`) and single-patient reports (Result 2).”

##### Box 2: Example diagrammatic representation

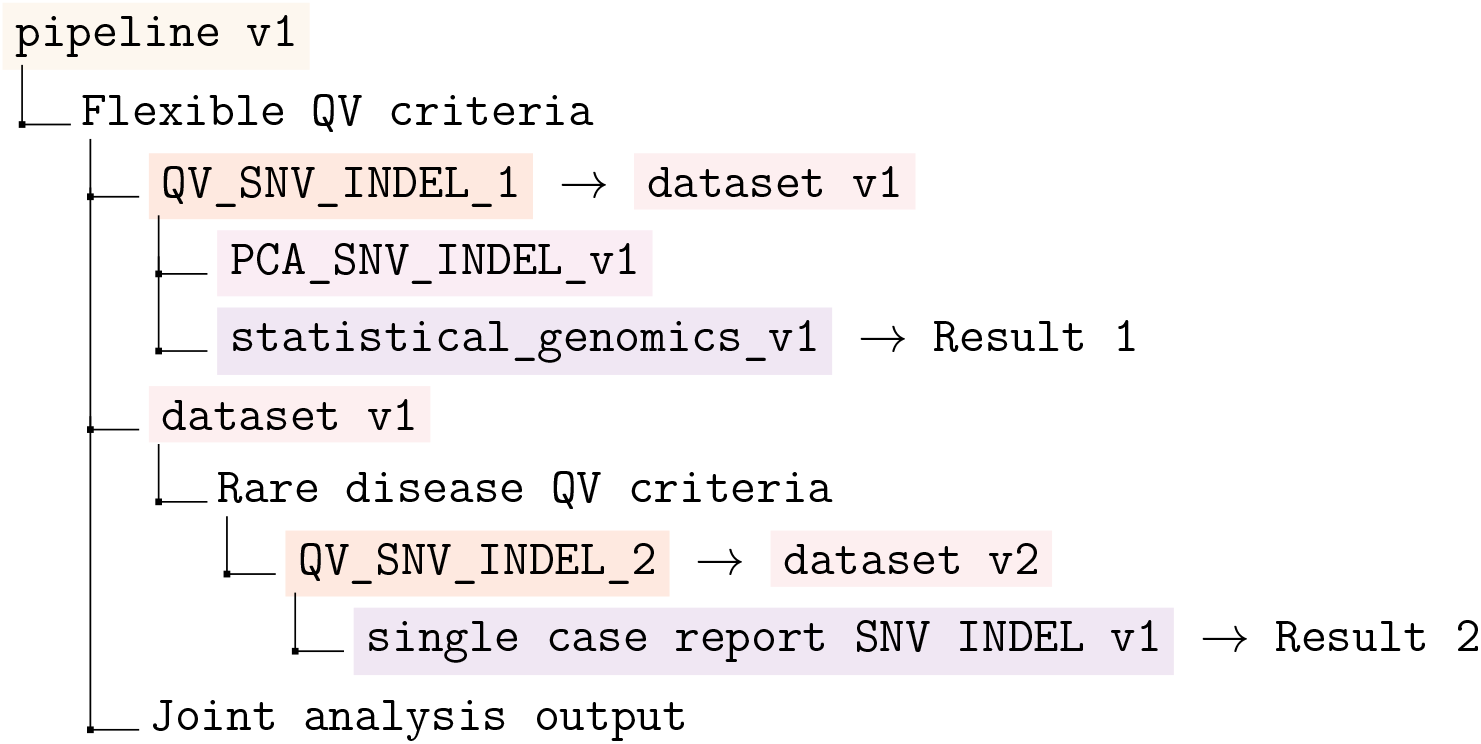

Joint analysis output from:

Result 1 = Cohort-level association signal (e.g. variant P-value).

Result 2 = Single variant report per patient.

### 2.2 Usage in a rare disease cohort validation study

We validated the QV framework on an in-house rare disease cohort of 940 individuals using Whole Exome Sequencing (WES) comparing a conventional manual implementation with a QV-based YAML configuration. The analysis targeted rare variants (Minor Allele Frequency (MAF) *<* 0.01) in known disease genes from the Genomics England “Primary immunodeficiency or monogenic inflammatory bowel disease” panel, retrieved via PanelAppRex (25). This yielded 6,026 candidate variants annotated with 376 information sources, prepared in R using the GuRu variant interpretation tool and imported from gVCF files processed by Variant Effect Predictor (VEP).

We applied the first eight American College of Medical Genetics and Genomics (ACMG) criteria for pathogenicity scoring (1), six of which were relevant to this cohort. The manual pipeline encoded each criterion directly, while the QV workflow read the same definitions from a YAML file. The YAML criteria included `ACMG_PS1` (known pathogenic amino acid change), `ACMG_PS3` (supporting functional evidence), `ACMG_PS5` (compound heterozygosity), and frequency- and segregation-based criteria (`PM2, PM3`). Criteria `PS2` and `PS4` were not applicable in this cohort.

### 2.3 Usage in a GWAS validation study

We next applied the QV criteria framework to a GWAS using HapMap3 Phase 3 (R3) consensus genotypes on 1397 individuals (26). Again, two pipelines were executed with identical inputs and parameters: one hard-coded and one driven by the QV file. This QV set defined common GWAS thresholds: restriction to autosomal, biallelic SNPs; minimum sample call rate of 95%; variant call rate of 95%; minor allele frequency *≥* 1%; and Hardy–Weinberg equilibrium *p ≥* 1 *×* 10^*−*6^. After quality control, variants were LD-pruned and principal components (PC1–PC10) were computed, with sex included as an additional covariate. Logistic regression under an additive model was then performed with a binary simulated phenotype using PLINK. The outputs of the two pipelines were captured and compared across each main PLINK stage. Manhattan plots, Principal Component Analysis (PCA) plots, and md5 checksums were used to confirm exact reproducibility between the hard-coded and QV-driven analyses.

For benchmarking, Message-Digest Algorithm 5 (MD5) checksums were uniquely reported for the GWAS study because PLINK output files are exactly reproducible between runs. In contrast, VCF files used in the other validation studies include variable header fields such as BCFtools view command with a timestamp, which changes with each run and alters the MD5 value. For those cases, we instead report variant count and content.

### 2.4 Usage in a WGS validation study with GIAB and Exomiser

We next applied the QV framework to a WGS trio analysis using the Genome In A Bottle Chinese Trio (HG005-HG007, PRJNA200694, GRCh38 v4.2.1) of the National Institute of Standards and Technology (27). Two pipeline phases were executed with identical inputs and parameters: one hard-coded and one driven by the QV file. Both phases applied identical QC and study filters and included a gene-panel style analysis using the paediatric disorders panel (panel 486; 3,853 genes (25)). The upstream processing used BCFtools for region restriction using BED overlap, site-level thresholds on QUAL and INFO/DP (using computed site depth from per-sample FORMAT/DP when absent), and per-sample thresholds on FORMAT/DP and FORMAT/GQ with exclusion of missing genotypes. Composite criteria were applied to require either all samples to pass or at least one sample to pass. The downstream filtered trio VCF was analysed with Exomiser using the same trio.ped input and without using Human Phenotype Ontology (HPO) terms.

## 3 Results

### 3.1 Validation rare disease cohort case study

We validated the QV framework using WES analysis with ACMG-based criteria on a rare disease cohort of 940 individuals, comparing a conventional pipeline with parameters defined internally (QV manual) to the new external YAML-based implementation (QV yaml). As shown in **Figure S1**, the outputs from both methods were identical, demonstrating a 100% match. This confirmed that our framework of a standalone, shareable, QV criteria file can be imported and applied programmatically with equivalent accuracy, providing a reproducible resource that is adaptable across different pipelines and programming environments.

### 3.2 Validation in a common variant GWAS

To demonstrate the integration of the QV framework with established best practices in GWAS (28), we validated it in a standard HapMap3 Phase 3 GWAS by again running two equivalent analyses: a conventional pipeline with parameters defined internally and a YAML-based implementation that externalised all settings. As shown in **Figure S2**, the Manhattan and PCA plots were identical between the two methods, and the MD5 checksums of all PLINK outputs matched exactly. These results confirm that QV parameterisation reproduces the original workflow precisely while improving clarity, transparency, and reusability.

### 3.3 Validation in a WGS study with GIAB and Exomiser

To demonstrate the ease and benefit of using QV parameterisation in established WGS analysis pipelines, we conducted a trio validation study using the Genome in a Bottle (GIAB) Chinese Trio (HG005-HG007, GRCh38 v4.2.1) and the Exomiser tool for variant annotation and interpretation (29). Two equivalent analyses were run: one with hard-coded thresholds and one using an external QV YAML file specifying the same parameters. Both applied identical QC and study filters and restricted analysis to the PanelAppRex paediatric disorders panel (3,853 genes). Results were identical: variant counts matched at each step, and Exomiser outputs produced the same candidate genes and variants. **Figure S3** shows this agreement. This validation confirms that a shareable QV file reproduces the full variant interpretation workflow exactly, while aligning with established variant effect predictors and interpretation tools (29–31). Benchmarking showed that QV files introduce no computational overhead and scale equivalently to conventional implementations (**Supplemental 10.2, Figure S4**).

### 3.4 Implications

#### General applicability and reproducibility

Across validation studies, the QV framework reproduced conventional workflows in which parameters are embedded within scripts, while externalising those same variables into a portable, shareable format. The framework itself performs no filtering, calling, annotation, or interpretation, but provides a machine-readable layer for defining and reusing the qualifying variables that underpin these analyses. It complements tools such as GATK and BCFtools for processing, Ensembl VEP, SnpEff, FAVOR, and WGSA for variant effect prediction (30), and Exomiser and VarFish for interpretation (29; 31), by making their analytic criteria explicit.

#### Scalability and interoperability with genomic tools

The validation studies, covering clinical interpretation, genome-wide association analysis, and WGS trio interpretation, demonstrate that the QV framework generalises across distinct genomic contexts without altering analytical outcomes or adding computational overhead. The format further allows users to define, combine, and extend their own QV sets using simple declarative syntax, providing a scalable approach for reproducible genomics.

#### Traceability and confirmation of applied clinical standards

Each QV file includes a persistent identifier and checksum that can be stored in Electronic Health Record (EHR) or laboratory systems such as EPIC, Cerner, Clinisys, or REDCap. This links each patient’s analysis (including any associated PPIE input) to the exact QV set used, enabling transparent, auditable, and FAIR-compliant reporting. A clinician or molecular pathologist viewing a result in EPIC or Cerner can access the linked `qv_set_id` to verify the applied standards and filtering criteria. Automated genomic reports should include these details by default, ensuring full traceability without requiring access to the pipeline. For example, if a patient asks whether their genome was screened for breast cancer due to variants in *BRCA1* or *BRCA2*, the EHR-linked report referencing “qv acmg sf v3.3 20250828.json” confirms that the ACMG secondary findings guideline (v3.3) (32) was applied, including its defined gene set, thresholds, version, and standard.

## 4 Summary

This paper introduces a framework for integrating qualifying variants into genomic analysis pipelines, enhancing reproducibility, interpretability and the seamless translation of research findings into clinical practice.

## Data Availability

All data produced in the present work are contained in the manuscript

https://doi.org/10.5281/zenodo.17414191

https://github.com/DylanLawless/qvApp2025lawless

## Acronyms

ACMG: American College of Medical Genetics and Genomics
EHR: Electronic Health Record
FAIR: Findable, Accessible, Interoperable, and Reusable
GIAB: Genome in a Bottle
GWAS: Genome-Wide Association Study
HPO: Human Phenotype Ontology
MAF: Minor Allele Frequency
MD5: Message-Digest Algorithm
PCA: Principal Component Analysis
PPIE: Public and Patient Involvement and Engagement
PRS: Polygenic Risk Score
QC: Quality Control
QV: Qualifying Variant
RDF: Resource Description Framework
SNOMED CT: Systematized Nomenclature of Medicine-Clinical Terms
VCF: Variant Call Format
VEP: Variant Effect Predictor
WES: Whole Exome Sequencing
WGS: Whole Genome Sequencing

## 5 Funding

This project was supported through the grant Swiss National Science Foundation 320030_201060, and NDS-2021-911 (SwissPedHealth) from the Swiss Personalized Health Network and the Strategic Focal Area ‘Personalized Health and Related Technologies’ of the ETH Domain (Swiss Federal Institutes of Technology).

## 6 Acknowledgements

Acknowledgements We would like to thank all the patients and families who have been providing advice on SwissPedHealth and its projects, as well as the clinical and research teams at the participating institutions.

## 7 Contributions

DL designed the work and contributed to the manuscript. AS, SB, VS, DH, SÖ, JA, SF contributed to the manuscript. LJS and JF supervised the work, manuscript, and applied for funding.

## 8 Competing interests

The authors declare no competing interests.

## 9 Ethics statement

The projects were approved by the respective ethics committees of all participating centers (Cantonal Ethics Committee Bern, approval number KEK-029/11) and the study was conducted in accordance with the Declaration of Helsinki.

## 10 Supplemental

**Application of qualifying variants for genomic analysis**.

### 10.1 Validation study figures

**Figure S1:**
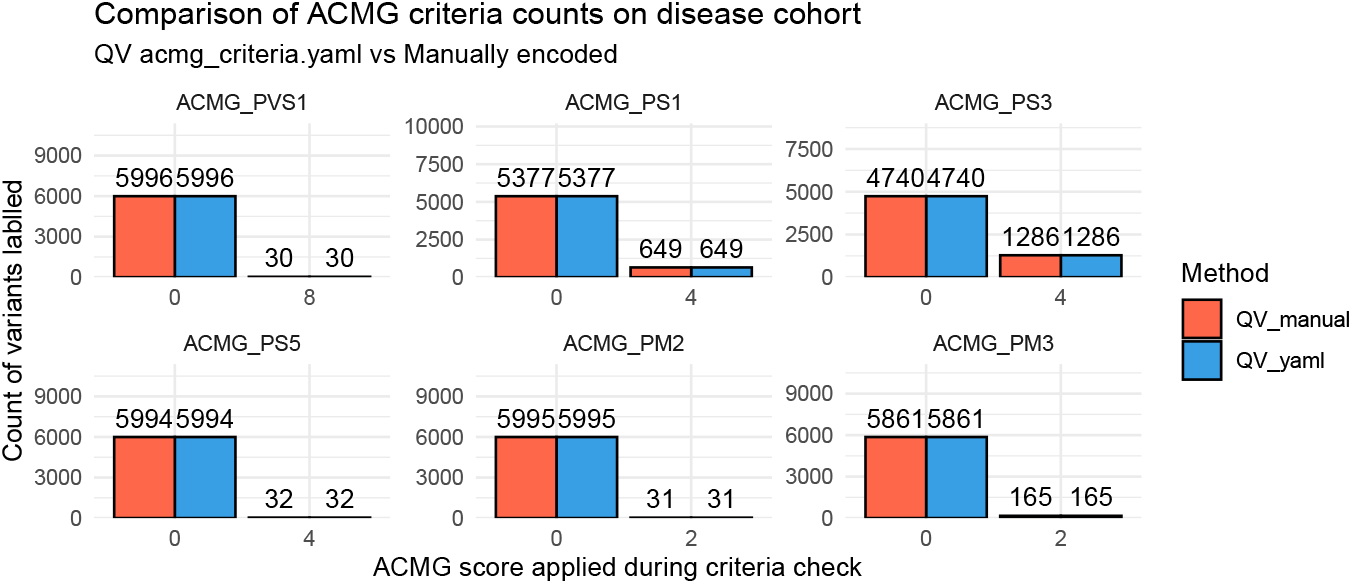
Validation case study of a rare disease cohort of 940 WES individuals using an ACMG criteria subset, demonstrating a 100% match between manually encoded and standalone YAML-based QV for assigning pathogenicity scores.

**Figure S2:**
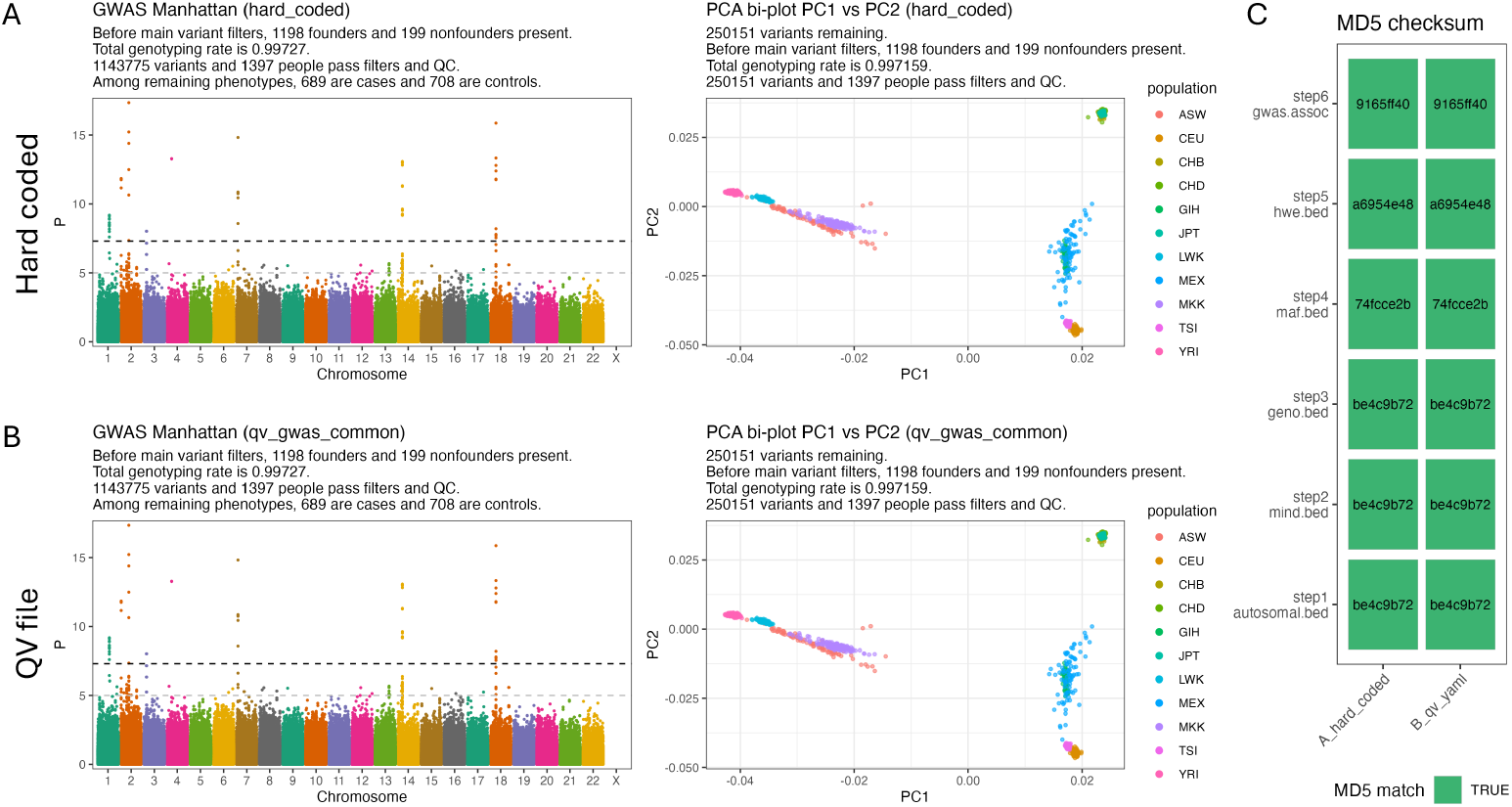
Validation in GWAS using QV parameterisation. (A) GWAS of simulated binary phenotypes in HapMap3 Phase 3 (R3) using a traditional variable embedded pipeline. Shown are the Manhattan plot of logistic regression results (left) and correction for population structure with principal component analysis (PC1 vs PC2, right). (B) Identical GWAS using a QV YAML configuration file. The Manhattan and PCA results are indistinguishable from panel A. (C) Verification of reproducibility. MD5 checksums of the main PLINK outputs are identical between panels A and B. The steps included processing of autosomal biallelic SNPs, sample call rate, variant call rate, minor allele frequency, Hardy–Weinberg equilibrium, and association results. The QV file encoded these thresholds (sample call rate ≥95%, variant call rate ≥95%, MAF ≥1%, HWE p ≥1e-6, autosomal biallelic SNPs only) together with covariates (sex and PC1-PC10) and logistic regression settings. This confirms that a shareable QV file reproduces hard-coded pipelines exactly while improving transparency and reusability.

**Figure S3:**
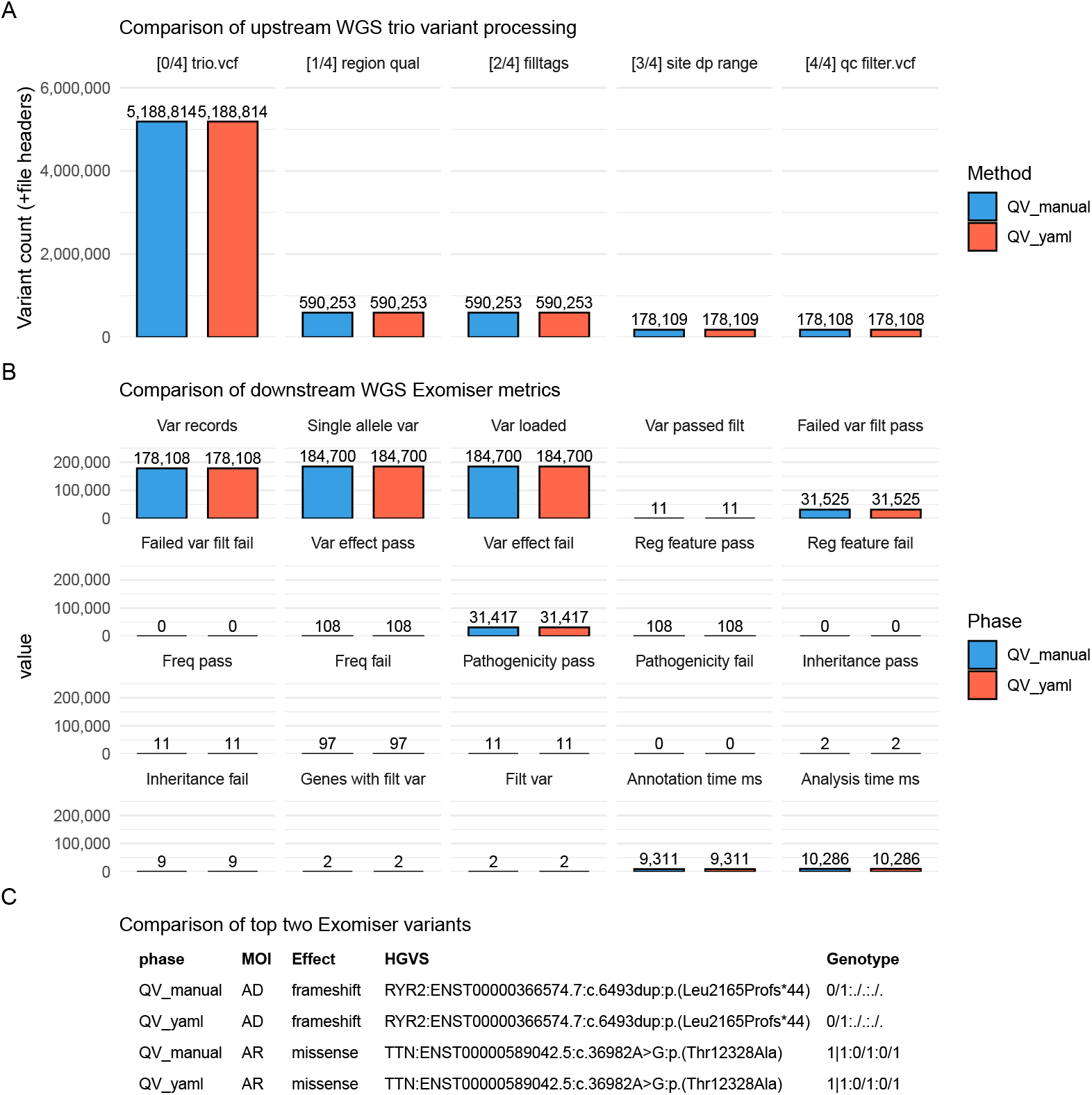
Validation of the trio Exomiser pipeline using QV parameterisation. **(A)** summarises upstream processing counts by file, **(B)** compares downstream Exomiser metrics, and **(C)** shows the key variant fields for the two variants identified. Variant counts in all panels confirm that intermediate files and final outputs are identical between configurations. The five preprocessing stages shown in (A) are: (0) input trio VCF, (1) gene panel region and quality filtering, (2) tag annotation, (3) site-level depth range filtering, and (4) final QC-filtered VCF. MOI, mode of inheritance; HGVS, Human Genome Variation Society nomenclature.

### 10.2 Computational benchmark

Runtime performance was equivalent between traditional and QV-based pipelines, as both read identical parameters from different sources. In the WGS trio validation study, pre-processing steps including filtering, QC, and gene panel selection completed in 16–17 seconds, with a median difference of ∼0.5 seconds favouring the QV YAML pipeline (**Figure S4**). An incidental one-off 5 second delay arose from Singularity initialisation for the yq utility (step 0), a system-specific effect on our HPC and unrelated to the framework itself.

**Figure S4:**
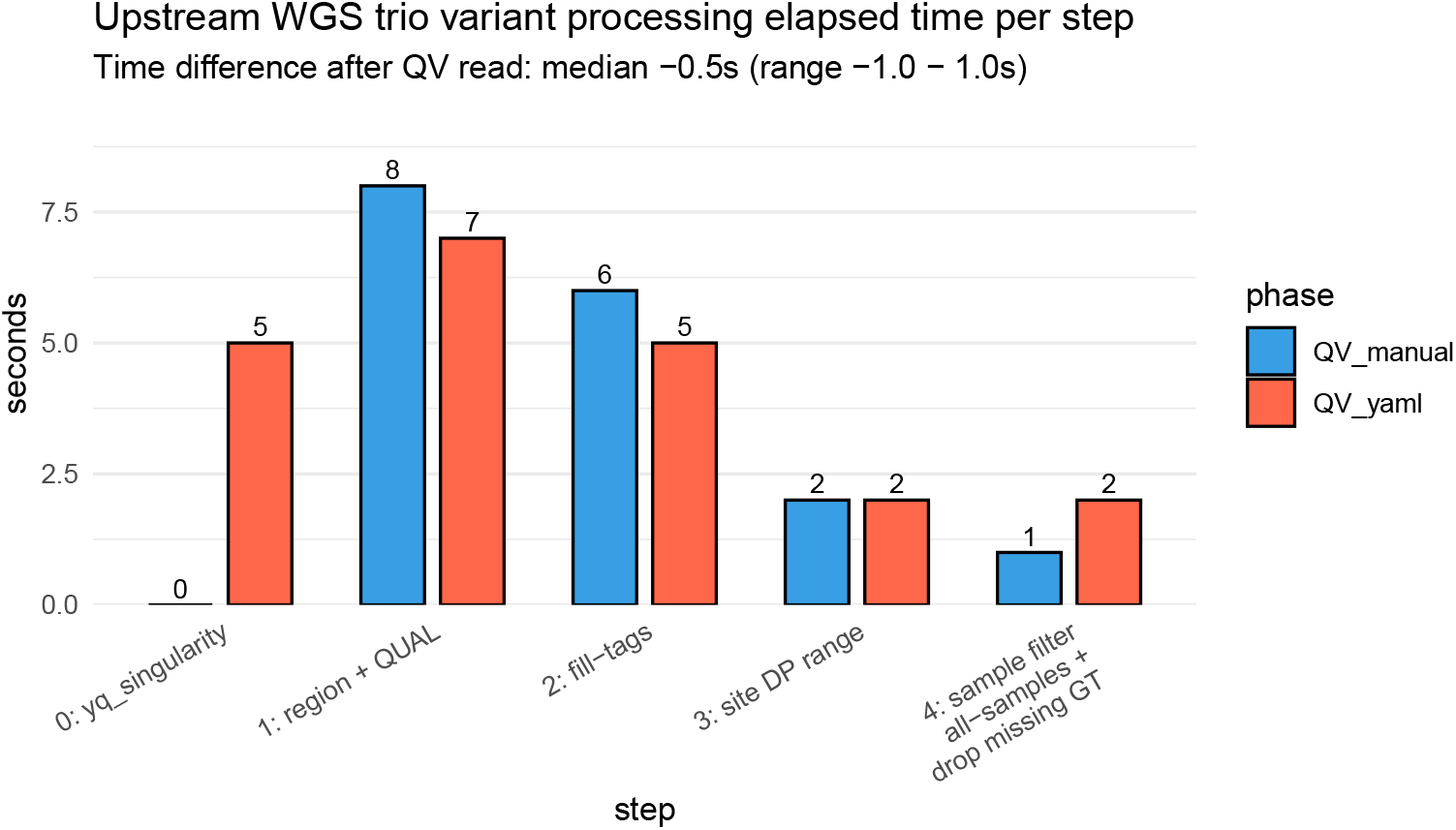
Benchmark of upstream preprocessing times in the WGS trio pipeline comparing QV-based and traditional (manually parameterised) configurations. Stepwise elapsed times were nearly identical across both methods (median difference ∼0.5 s), with a fixed 5 s overhead from optional Singularity initialisation of yq in the QV pipeline. The four preprocessing steps correspond to: (1) gene panel region and quality filtering of the trio VCF, (2) annotation of variant tags, (3) site-level depth range filtering, and (4) per-sample genotype filtering and exclusion of missing genotypes. All steps used `BCFtools` on VCF preprocessing, as illustrated in **Figure S3 (A)**.

### 10.3 How to build a QV file

We recommend YAML or JSON for portability and adoption. You can build a QV in three ways:

#### Option 1: use the HTML QV builder (Zenodo)

1. Open the HTML builder from the Zenodo repository.
2. Enter simple `key=value` statements in the left pane.
3. Copy or download the generated YAML.

Example input lines:

~~~
meta qv_set_id=“qv_gwas_common_v1_20250827”
meta version=“1.0.0”
meta title=“GWAS common QC”
meta authors=Alice,Bob
meta tags=GWAS,QC,PCA
filter maf_minimum field=MAF operator=“>=“ value=0.01 desc=“Minimum MAF”
filter hwe field=HWE_P operator=“>=“ value=1e-6 logic=keep_if
filter region_include desc=“include panel” field=OVERLAP(targets.exome.bed)
    >>>> operator=“>=“ value=1 logic=keep_if
criteria disease_panel logic=and desc=“HIGH impact within panel”
criteria disease_panel field=IMPACT operator=“==“ value=HIGH
criteria disease_panel field=OVERLAP(targets.exome.bed) operator=“>=“ value=1
meta description_patient=
    >>>> “There is a strong family history of early heart attacks.”
meta description_ppie=
    >>>> “The PPIE group reviewed and approved the criteria on 2025-08-15.”
~~~

#### Option 2: write YAML by hand

Minimal pattern:

~~~
meta:
  qv_set_id: qv_disease_panel_v1_20250828
  version: 1.0.0
  title: Disease panel
filter filters:
  region_include:
    description: Restrict to curated disease gene panel
    logic: keep_if
    field: OVERLAP(targets.disease_panel.bed)
    operator: “>=“
    value: 1
criteria:
  pathogenic:
     description: Variant classified as pathogenic or likely pathogenic
     logic: and
     conditions:
      -group: any_of:start
      -{ field: CLASS, operator: “==“, value: P }
      -{ field: CLASS, operator: “==“, value: LP }
      -group: any_of:end
notes:
   - Gene panel file defines the target regions
~~~

#### Option 3: write JSON

JSON equivalent of the minimal example:

~~~
{
   “meta”: {
       “qv_set_id”: “qv_disease_panel_v1_20250828”,
       “version”: “1.0.0”,
       “title”: “Disease panel filter”
   },
   “filters”: {
      “region_include”: {
        “description”: “Restrict to curated disease gene panel”,
        “logic”: “keep_if”,
        “field”: “OVERLAP(targets.disease_panel.bed)”,
        “operator”: “>=“,
        “value”: 1
      }
   },
   “criteria”: {
     “pathogenic”: {
       “description”: “Variant classified as pathogenic or likely pathogenic”,
       “logic”: “and”,
       “conditions”: [
           { “group”: “any_of:start” },
           { “field”: “CLASS”, “operator”: “==“, “value”: “P” },
           { “field”: “CLASS”, “operator”: “==“, “value”: “LP” },
           { “group”: “any_of:end” }
       ]
    }
 },
“notes”: [
    “Gene panel file defines the target regions”
  ]
}
~~~

#### Checksum and register

Record the checksum and register the release:

~~~
sha256sum qv/examples/qv_disease_panel_v1_20250828.yaml
# qv/registry/releases.csv
qv_set_id, version, checksum, file, date
qv_disease_panel_v1_20250828,1.0.0, ef6cf810b994…,
    > qv_disease_panel_v1_20250828.yaml, 2025-08-28
~~~

#### Versioning and IDs

Use a stable `qv_set_id` plus semantic version. Update the version on any change that affects selection or interpretation. Keep one file per release and never mutate published files.

#### Use in a workflow

Point your pipeline to the QV file:

~~~
# workflows/…/config.yaml
qv_file: “…/qv/registry/qv_disease_panel_v1_20250828.yaml”
~~~

It can be read programmatically at runtime, for example using yq in shell-based workflows or `yaml::read_yaml()` in R, providing the same parameters that would otherwise be embedded within pipeline configurations.

## Notes

### Competing Interest Statement

The authors have declared no competing interest.

### Funding Statement

This project was supported through the grant Swiss National Science Foundation (SNF) 320030_201060, and NDS-2021-911 (SwissPedHealth) from the Swiss Personalized Health Network and the Strategic Focal Area 'Personalized Health and Related Technologies' of the ETH Domain (Swiss Federal Institutes of Technology).

### Author Declarations

Summary statistics were used from studies which have been previously reported and approved by the respective ethics committees of all participating centers (Cantonal Ethics Committee Bern, approval number KEK-029/11) and the study was conducted in accordance with the Declaration of Helsinki.

### Summary of Updates

We reformatted the method implementation for clarity and added new validation studies to demonstrate usage and results: - A WGS trio analysis using the Genome In A Bottle Chinese Trio (HG005-HG007, PRJNA200694, GRCh38 v4.2.1). - A GWAS using HapMap3 Phase 3 (R3) consensus genotypes on 1397 individuals.

